# Beyond Agreement: a real-world study of the workflow gap between echocardiography and timely structural cardiac assessment How a Validation Study Exposed a Hidden Gap in Cardiac Care

**DOI:** 10.64898/2026.05.12.26352129

**Authors:** Marta Afonso Nogueira, Fernanda Costa Ferreira, Élia Batista, Sara Eira, Gonçalo Proença, Carla Matias, Istvan Kecskes

## Abstract

**Objectives:** To assess agreement between Cardio-HART (CHART) and echocardiography for left ventricular ejection fraction (LVEF) estimation and heart failure (HF) classification in a real-world predominantly ischaemic cohort, while examining whether a point-of-care structural and functional assessment tool could reveal a broader workflow gap between the nominal availability of echocardiography and timely cardiac assessment in routine care.

**Design:** Prospective single-centre cohort study.

**Setting:** Secondary-care cardiology service at Cascais Hospital, Lisbon, Portugal.

**Participants:** Forty-seven adults referred for cardiology evaluation with suspected HF or followed in a hospital HF clinic.

**Primary and secondary outcome measures:** Agreement between CHART-derived and echocardiographic LVEF by Bland-Altman analysis; diagnostic performance for HF phenotypes; comparison with the Teichholz method.

**Results:** Mean age was 65.6±15.9 years; 78.7% of participants had HF and 43.2% of HF cases were ischaemic. CHART showed a mean LVEF bias of +1.92% versus echocardiography, with 95% limits of agreement from -14.6% to +18.4% and a mean absolute error of 6.09%. Agreement was strongest in HF with reduced ejection fraction (HFrEF) and HF with mildly reduced ejection fraction (HFmrEF), and lower in HF with preserved ejection fraction (HFpEF). Diagnostic area under the curve for HFrEF classification was 0.89. Compared with the Teichholz method, CHART showed a lower root mean square error relative to Simpson’s biplane LVEF.

**Conclusions:** CHART showed clinically credible performance for LVEF estimation and HF stratification, particularly in reduced-EF phenotypes. However, the most important finding of this study was not agreement alone. By performing credibly in a cardiology-based real-world setting, CHART exposed a previously under-recognised workflow gap between the nominal availability of echocardiography and timely access to structural cardiac assessment in routine care. The study therefore suggests that the value of CHART lies not only in diagnostic performance, but in making visible, and potentially narrowing, a hidden but consequential gap in cardiac assessment pathways. Larger studies are warranted, particularly for HFpEF and across broader clinical workflows.

**Strengths and limitations of this study:**

- This was a prospective real-world study conducted in a cardiology-led secondary-care setting.
- CHART was evaluated against same-day echocardiography in almost all participants, using Simpson’s biplane LVEF as the reference standard.
- The study contributes not only agreement data, but also operational insight into the gap between the assumed presence of echocardiography and its timely use in real clinical workflows.
- The sample size was small, limiting precision and subgroup interpretation.
- HFpEF findings should be regarded as exploratory and require confirmation in larger multicentre cohorts.

## Introduction

Heart failure (HF) is a major and growing global health problem, affecting more than 64 million people worldwide and placing a substantial burden on patients, health systems and healthcare expenditure.^1^ HF is frequently under-recognised in both acute and non-acute settings because its symptoms may be non-specific, overlap with other disorders, or develop gradually over time.^2–4^ These diagnostic challenges are especially pronounced in HF with preserved ejection fraction (HFpEF), in which symptoms may be subtle and conventional clinical assessment often lacks specificity.^3,4^

Although biomarkers such as B-type natriuretic peptide and N-terminal pro-B-type natriuretic peptide can support case finding, they do not provide structural or functional cardiac assessment.^5,6^ Echocardiography therefore remains the gold standard for assessing left ventricular ejection fraction (LVEF), defining cardiac structure and function, and guiding HF diagnosis, phenotyping, follow-up and management.^7,12^ Yet the central problem in real-world care is often described too narrowly as one of limited echocardiography availability in primary care or resource-limited settings. That description is incomplete.

In practice, the more important limitation is frequently the gap between the theoretical availability of echocardiography and its timely use within the workflow in which decisions are actually being made. Echocardiography may exist within the institution and still remain unavailable at the point of need because access depends on acquisition capacity, reporting capacity, specialist review, pathway sequencing and prioritisation. Under these circumstances, clinicians often rely on symptoms, biomarkers and ECG while awaiting imaging, even though ECG alone cannot provide the structural and functional information required for LVEF assessment, HF phenotyping, valve evaluation or many causes of clinical deterioration.^5,8,12^

This distinction matters. The real clinical gap is not simply between a hospital that has echocardiography and one that does not. It is the gap between gold-standard capability and real-world access to timely structural cardiac assessment. That gap may affect not only primary care, but also specialist and hospital workflows in which cardiovascular decisions must still be made without immediate structural information. It is easily normalised because the gold standard is assumed to be available somewhere in the system, even when it is not practically accessible when and where needed.

This issue is particularly relevant in high-risk populations such as patients with coronary artery disease (CAD), in whom regional wall motion abnormalities, progressive systolic dysfunction and evolving HF may require timely reassessment.^9^ Such patients often have multiple comorbidities, including diabetes, chronic kidney disease and obesity, which can obscure the clinical picture and delay recognition of HF. In these contexts, delayed access to echocardiography may not merely slow confirmation of disease; it may delay the point at which structural and functional deterioration becomes visible at all.

Cardio-HART (CHART) is a next-generation ECG-based diagnostic platform that combines advanced biosignal analysis, 12-lead ECG and artificial intelligence to support earlier detection and triage of cardiovascular disease, including HF and valvular disease.^10^ Previous work has shown promising diagnostic performance across diverse HF presentations, particularly in cohorts enriched for structural, diastolic and valvular abnormalities.^11^ However, typical ischaemic HF, commonly encountered in hospital cardiology practice, was underrepresented in prior validation.

The present study was therefore designed to assess agreement between CHART and echocardiography for LVEF estimation and HF classification in a real-world predominantly ischaemic cohort. However, what began as a technical agreement study proved more revealing than expected. By testing a rapid point-of-care tool in a cardiology-led environment in which echocardiography was assumed to be readily available, the study also provided an opportunity to observe whether a clinically meaningful workflow gap remained hidden within routine practice itself. In that sense, if CHART performed credibly in this setting, it could do more than agree with echocardiography: it could expose the extent to which timely structural cardiac assessment was still missing from workflows presumed already to contain it.

## Materials and methods

### Study design and population

This was a prospective single-centre cohort study conducted between January 2022 and July 2023 at Cascais Hospital, Lisbon, Portugal. Patients were included if they met one of the following criteria: (1) referral for cardiology evaluation with suspected HF, based on symptoms, elevated natriuretic peptides and/or abnormal echocardiographic findings from an external facility; or (2) follow-up in the hospital HF clinic. Patients with poor echocardiographic windows or acute decompensated HF were excluded. European Society of Cardiology guidelines were followed for the diagnosis and treatment of acute and chronic HF.^12^ Approval was obtained from the hospital ethics committee, and all patients provided informed consent before enrolment. Participant selection and study flow are summarised in Figure 1.

**Figure 1.**
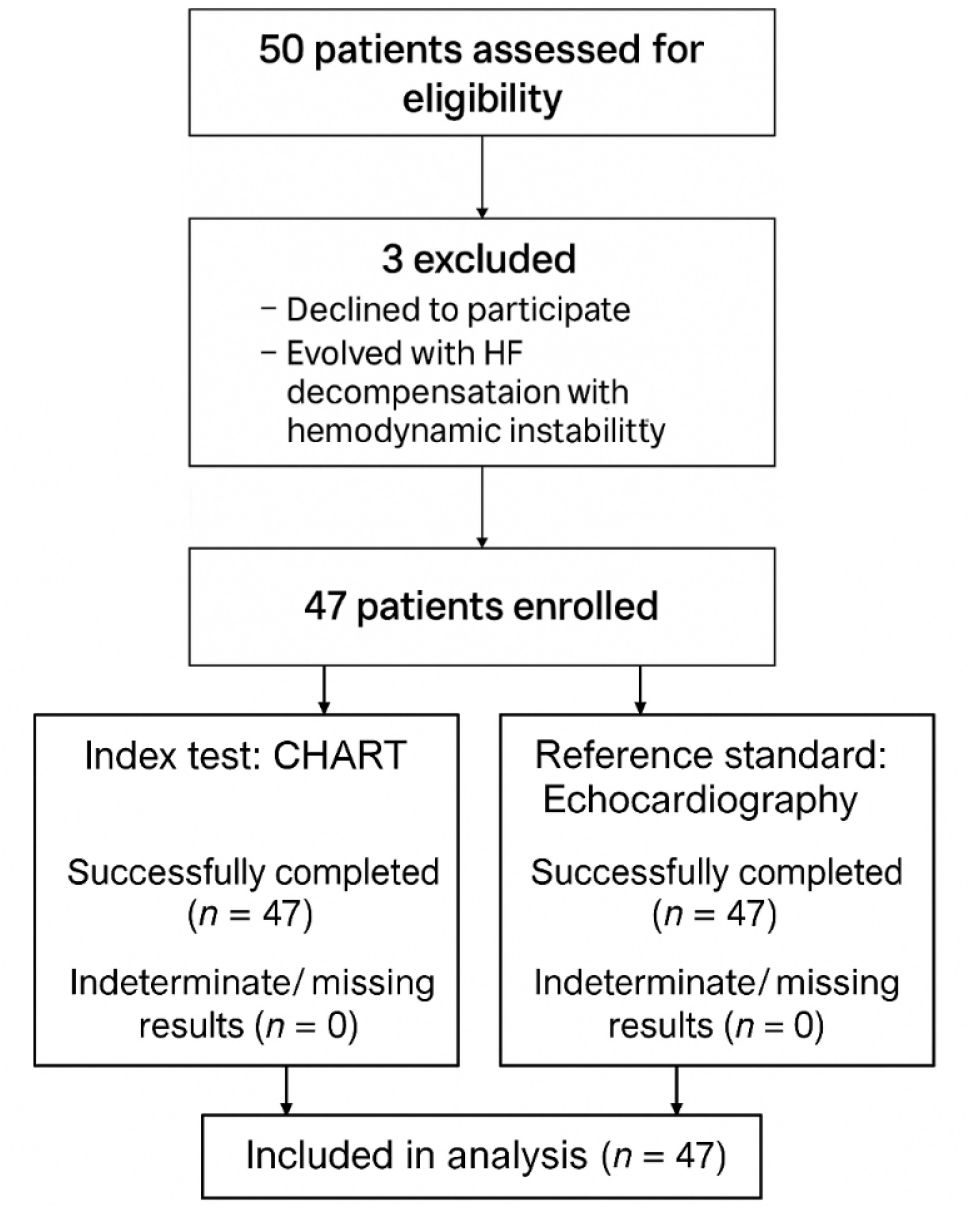
Participant flow diagram.

### Echocardiographic assessment

Standard transthoracic echocardiography (TTE) was performed by cardiopulmonary technicians and reviewed by cardiologists, following European Association of Cardiovascular Imaging recommendations for image acquisition.^13^ The echocardiography laboratory held EACVI accreditation for TTE. LVEF derived by Simpson’s biplane method was used as the reference standard.

Additional echocardiographic variables recorded included body surface area, global longitudinal strain, E/A ratio, E/e^′^ ratio, left ventricular outflow tract velocity-time integral, left ventricular internal dimension in diastole, indexed left ventricular mass, Teichholz-derived ejection fraction, relative wall thickness, deceleration time, aortic valve maximum velocity, mitral valve area, pressure half-time and pulmonary artery systolic pressure.

### CHART assessment

CHART assessment was performed on the same day as echocardiography in all but one patient, who underwent CHART the following day. Operators completed a 6-hour training course with competency assessment. Cardiologists reviewing echocardiograms and cardiopulmonary technicians performing CHART were blinded to CHART results.

### Performance evaluation

Diagnostic performance was evaluated using common binary performance metrics. In addition to sensitivity and specificity, positive predictive value, negative predictive value, F1-score, Cohen’s kappa and SEsp90% were calculated. Performance was assessed across three clinically relevant cut-offs: all HF, EF <50% and EF <40%. Definitions and descriptions of the performance metrics are provided in Supplementary Table 1.

### Statistical analysis

Agreement between CHART-derived LVEF and echocardiographic LVEF was assessed using Bland– Altman analysis, including mean bias and 95% limits of agreement (LoA). A difference of 15% was prespecified as clinically acceptable. Root mean square error (RMSE), area under the receiver operating characteristic curve (AUC) and binary classification measures were also calculated.

AUC was calculated for:

- HFpEF+ versus unlikely HF
- HFmrEF+ versus unlikely HF and HFpEF
- HFrEF versus all remaining groups

Binary classification using 50% as the LVEF threshold included sensitivity, specificity, accuracy and F1-score for EF <50%.

## Results

### Study population

Forty-seven patients were included. Mean age was 65.66±15.90 years and 70% were male. According to echocardiographic assessment, 78.7% had HF, of whom 64.9% had LVEF <50%. Structural heart disease was present in 70.2%, functional heart disease in 61.7%, wall motion abnormalities in 63.8%, valvular heart disease in 42.6% and atrial fibrillation in 23.4% (Table 1).

**Table 1.** Patients’ sociodemographic and clinical characteristics.

| | Mean ± SD/<br>n (%) | Proportion with<br>available data |
| --- | --- | --- |
| **Sociodemographic characteristics** | **n=47** |  |
| Age [years] | 65.66±15.90 | 100% |
| Male | 33 (70%) | 100% |
| Female | 14 (30%) | 100% |
| Height [cm] | 165.45±10.73 | 100% |
| Weight [kg] | 77.47±17.76 | 100% |
| **Clinical Characteristics, mean** | **n=47** |  |
| BSA [m<sup>2</sup>] | 1.87±0.25 | 100% |
| EF Biplane [%] | 49.99±12.31 | 100% |
| GLS [%] | -15.21±4.11 | 94% |
| E/A | 0.93±0.33 | 83% |
| E/e' Avg | 11.11±6.27 | 60% |
| LVOT VTI [cm] | 19.28±5.73 | 74% |
| LVIDd [mm] | 52.58±8.08 | 89% |
| LV Mass / ASC | 107.00±32.06 | 89% |
| EF Teich [%] | 44.05±15.98 | 89% |
| RWT [%] | 39.06±10.34 | 89% |
| DT [ms] | 243.11±106.59 | 89% |
| AV Vmax [m/s] | 1.66±0.63 | 87% |
| MVA by PHT [cm<sup>2</sup>] | 3.71±1.31 | 100% |
| PASP [mmHg] | 27.43±12.35 | 89% |
| **Echocardiographic-based characterization** | **n=47** |  |
| HF | 37 (78.7%) | 100% |
| Valvular Heart Disease | 20 (42.6%) | 100% |
| Structural Heart Disease | 33 (70.2%) | 100% |
| Functional Heart Disease | 29 (61.7%) | 100% |
| Wall Motion Abnormality (Ischaemic Disease) | 30 (63.8%) | 100% |
| Atrial Fibrillation | 11 (23.4%) | 100% |
| Low EF (EF<50%) | 24 (51.1%) | 100% |
| Pacemaker | 5 (10.6%) | 100% |
| **HF main source** | **n=37** |  |
| Ischaemic HF | 16 (43.2%) | 100% |
| Valvular HF | 9 (24.3%) | 100% |
| Structural HF | 8 (21.6%) | 100% |
| Right Ventricular or Pulmonary Hypertension HF | 3 (8.1%) | 100% |
| Diastolic HF (borderline case) | 1 (2.7%) | 100% |

BSA: body surface area; EF: ejection fraction; GLS: global longitudinal strain; LVOT VTI: left ventricular outflow tract velocity-time integral; LVIDd: left ventricular internal dimension in diastole; LV Mass/ASC: left ventricular mass index; RWT: relative wall thickness; DT: deceleration time; AV Vmax: aortic valve maximum velocity; MVA: Mitral valve area; PHT: pressure half-time; PASP: Pulmonary Artery Systolic Pressure.

Regarding HF aetiology, 43.2% of patients with HF had ischaemic HF, defined by wall motion abnormality on echocardiography. Among non-ischaemic HF cases, 24.3% were valvular, 21.6% structural, 8.1% right ventricular or pulmonary hypertension-related and 2.7% diastolic.

### CHART diagnostic performance

Using echocardiography as the reference standard, CHART achieved sensitivity of 86.5% and specificity of 80.0% for differentiating patients with HF from those unlikely to have HF. The corresponding F1-score was 90.1% (Table 2).

**Table 2.** Binary performance metrics of HF classification with different cutoffs.

| Properties of binary HF criteria |  |  |  |  | Performance of CHART HF |  |  |  |  |  |  |  |
| --- | --- | --- | --- | --- | --- | --- | --- | --- | --- | --- | --- | --- |
| Cutoff | Negative | Positive | Prev% | PR% | SE% | SP% | PPV% | NPV % | F1% | K% | SEsp90 % | ACC% |
| All HF | Unlikely | HFpEF<br>HFmrEF<br>HFrEF | 78.7 | 72.3 | 86.5 | 80.0 | 94.1 | 61.5 | 90.1 | 59.9 | 73.4 | 85.1 |
| EF<50 % | Unlikely<br>HFpEF | HFmrEF<br>HFrEF | 51.1 | 48.9 | 79.2 | 82.6 | 82.6 | 79.2 | 80.9 | 61.7 | 65.9 | 87.3 |
| EF<40 % | Unlikely<br>HFpEF<br>HFmrEF | HFrEF | 23.4 | 19.2 | 63.6 | 94.4 | 77.8 | 89.5 | 70.0 | 62.0 | 78.0 | 87.2 |

Prev: prevalence; PR: positive rate; SE: sensitivity; SP: specificity; PPV: positive predictive value; NPV: negative predictive value; F1: F1 score; K: Cohen's kappa; SEsp90: estimated sensitivity when the SP is fixed to 90% using parametric ROC curve; ACC: accuracy.

For distinguishing HFmrEF/HFrEF from unlikely HF/HFpEF, sensitivity was 79.2% and specificity was 82.6%. For distinguishing HFrEF from unlikely HF/HFpEF/HFmrEF, CHART achieved sensitivity of 63.6% and specificity of 94.4%.

The confusion matrix showed moderate capability for HF categorisation compared with the echocardiographic ground truth (Table 3). CHART performed best in identifying non-HF from HF and in detecting HFrEF, whereas HFpEF was the most difficult category to predict accurately. Compared with the conventional Teichholz method, CHART demonstrated comparable AUC values and a lower RMSE relative to Simpson’s biplane LVEF, indicating closer alignment with the reference standard (Table 4). CHART achieved an RMSE of 8.80%, whereas the Teichholz method showed an RMSE of 11.80%.

**Table 3.**
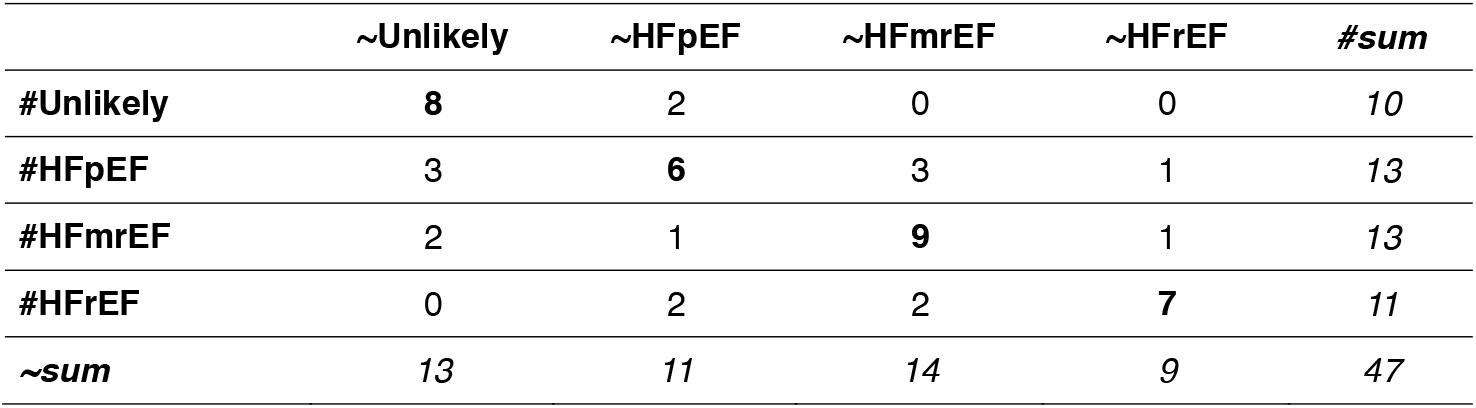
Confusion Matrix of Heart Failure Classification by CHART.

**Table 4.**
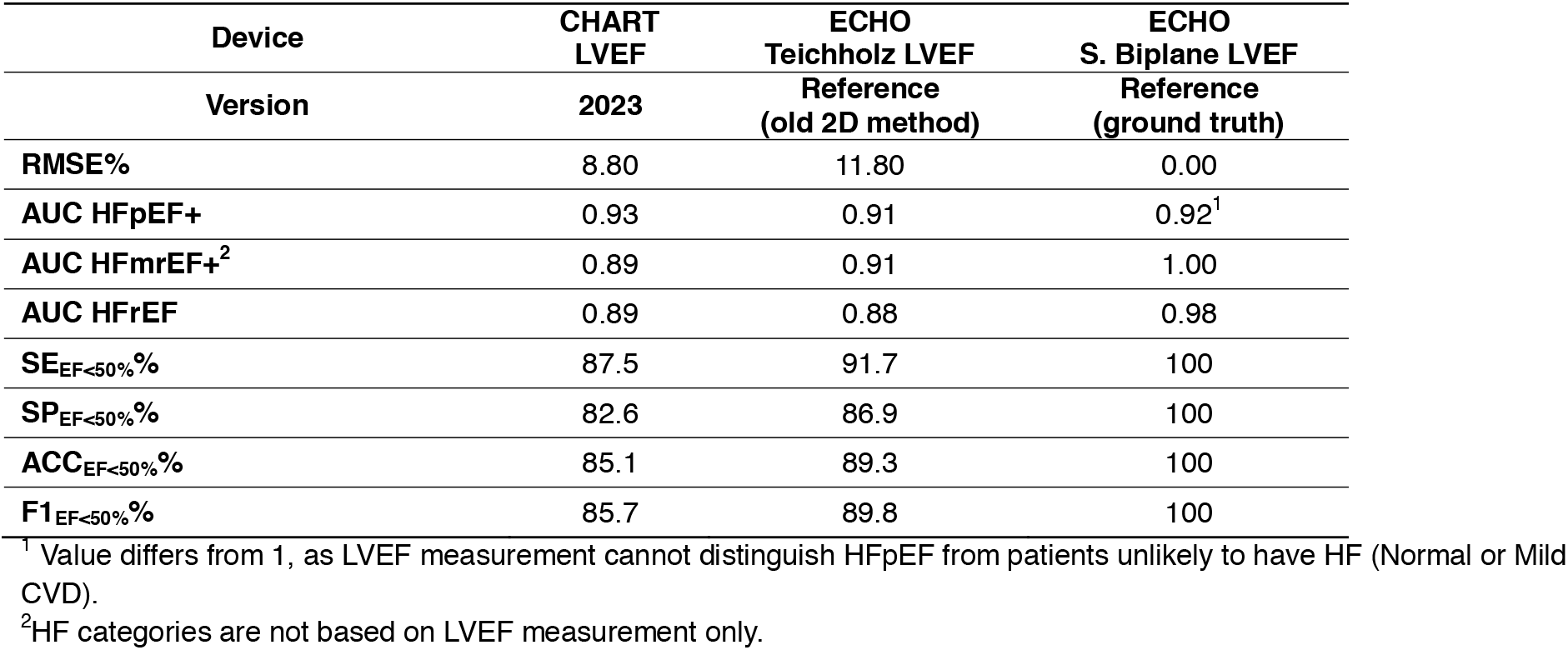
Performance comparison of CHART and reference method for estimating LVEF.

### Agreement between CHART and echocardiography

Bland–Altman analysis showed that CHART-derived LVEF had a mean bias of +1.92% versus echocardiography, with 95% LoA from −14.6% to +18.4% and a mean absolute error of 6.09%. By comparison, Teichholz-derived LVEF showed a mean bias of −3.8%, with wider LoA from −27.0% to +19.4% and a mean absolute error of 8.46%. Agreement between CHART and echocardiography is illustrated in Figure 2.

**Figure 2.**
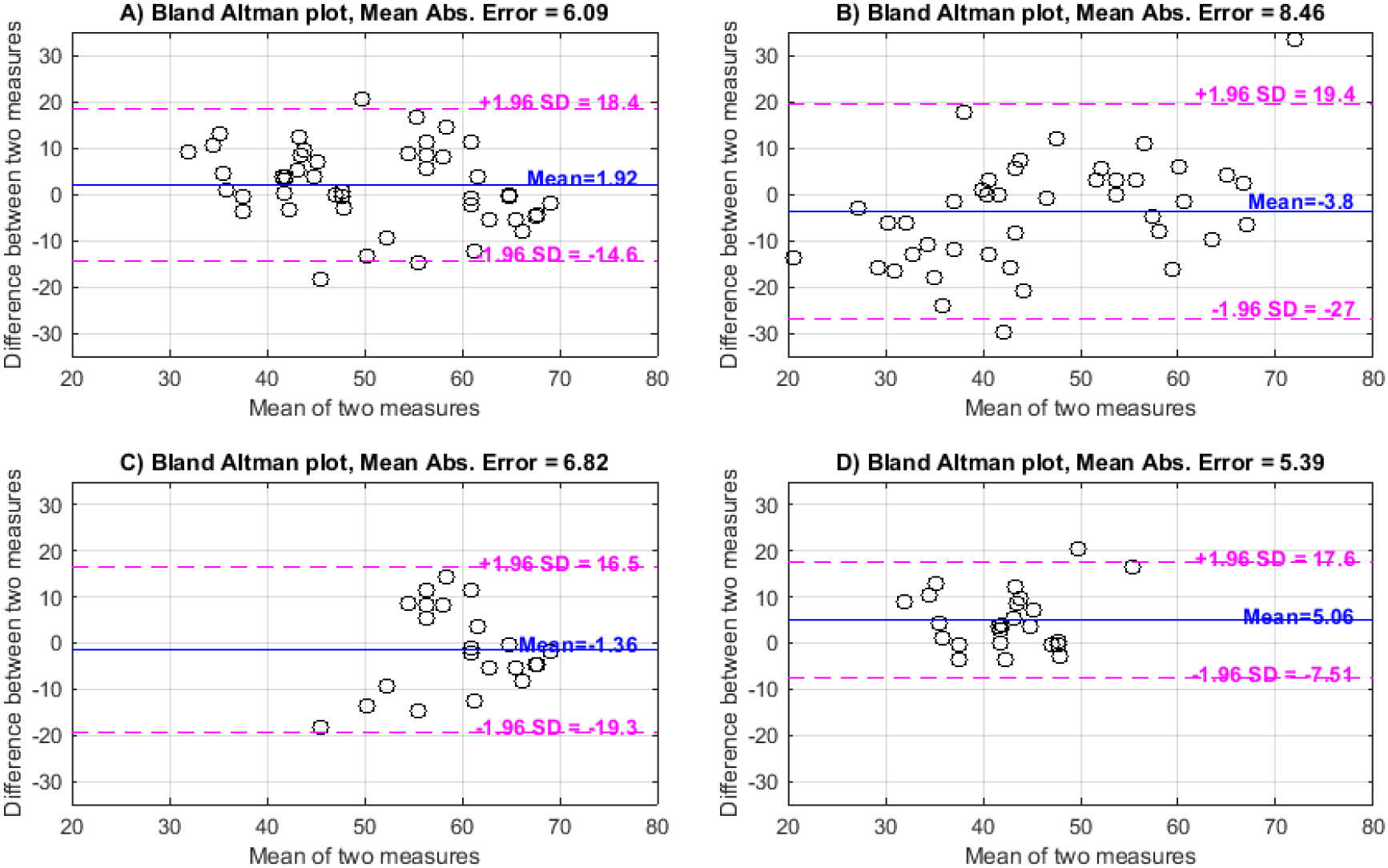
**A)** Bland-Altman plot of CHART LVEF with reference S. Biplane LVEF; **B)** Bland-Altman plot of Teichholz LVEF with reference S. Biplane LVEF; **C)** Bland-Altman plot of CHART LVEF including only patients with normal function and HFpEF patients; **D)** Bland-Altman plot of CHART LVEF including only HFmrEF and HFrEF patients.

In patients with normal or mildly impaired function (HFpEF), CHART showed a lower mean bias (−1.36%) but higher variability (LoA −19.3% to +16.5%; mean absolute error 6.82%). In patients with reduced LVEF (HFmrEF/HFrEF), CHART showed a higher mean bias (+5.06%) but lower variability (LoA −7.5% to +17.6%; mean absolute error 5.39%). Overall, agreement was stronger in reduced-EF phenotypes than in HFpEF.

The observed level of agreement is also consistent with previously reported variability between established imaging modalities for LVEF assessment, including echocardiography, cardiac MRI and nuclear imaging.^15–17^

## Discussion

This study was originally conceived as an agreement study: a real-world assessment of whether CHART could estimate LVEF and classify HF in a predominantly ischaemic cardiology cohort with sufficient credibility relative to echocardiography. On that objective, the findings were positive. CHART showed clinically credible agreement with Simpson’s biplane LVEF, lower RMSE than the Teichholz method, and strongest performance in reduced-EF phenotypes, particularly HFrEF and HFmrEF.

However, the most important finding of this study was not agreement alone.

Because CHART performed credibly at the point of care in a cardiology-led environment, it exposed something that had previously remained obscured within routine practice: a workflow failure hidden inside the apparent presence of the gold standard. The study did not simply confirm that CHART could be compared with echocardiography. It revealed that the distance between nominal echocardiographic availability and timely structural cardiac assessment was not trivial, but clinically meaningful, even in a specialist setting where that gap might reasonably have been assumed to be small.

That distinction matters. Much of the conventional literature frames delayed diagnosis as a consequence of limited access to echocardiography, particularly outside specialist care.^5,8^ While true, that framing risks understating the problem by treating availability as if it were equivalent to access. This study suggests otherwise. Echocardiography may be present in the environment and yet remain absent, particularly at the beginning of the workflow in which decisions are actually being made. When that happens, the deficit is easy to miss because the gold standard exists somewhere in the system and its absence at the point of care becomes normalised.

In that sense, the study widened its own scope. What began as a technical comparison unexpectedly laid bare the operational consequence of a system that assumes access where there may only be theoretical availability. The surprise was not merely that CHART performed reasonably well, but that its performance made visible how much structurally informative cardiac assessment had been deferred or delayed within routine care, despite the setting being cardiology-led.

This is especially relevant in an ischaemic cohort. Patients with coronary artery disease are at high risk of progressive systolic dysfunction and regional wall motion abnormalities, and may deteriorate structurally before that deterioration is adequately reflected in routine symptom-based or ECG-based follow-up.^9^ In such populations, the absence of timely structural assessment is not a benign workflow inconvenience. It has the potential to affect treatment timing, risk assessment and care prioritisation.

CHART performed best in detecting HF and HFrEF, while accuracy was lower in HFpEF. This pattern is clinically plausible. HFpEF remains challenging across diagnostic modalities, including echocardiography, and the small HFpEF subgroup in the present cohort limits firm inference.^12,18,19^ The wider LoA observed in HFpEF are therefore more appropriately interpreted as exploratory than as evidence of intrinsic technological inadequacy. By contrast, performance in reduced-EF phenotypes was stronger and more consistent, which reinforces the relevance of CHART in areas of care where delayed identification of impaired systolic function may have direct management implications.

The present findings should not be interpreted as suggesting that CHART replaces echocardiography. That is not the claim, and the data do not support interchangeability across all clinical circumstances. The more important implication is different. CHART appears capable of narrowing the otherwise hidden gap between gold-standard capability and real-world access to structural and functional cardiac information. Its significance lies not only in its diagnostic performance metrics, but in its ability to reveal and potentially reduce a workflow deficit that had become largely invisible.

This observation may have implications beyond HF clinics alone. Other hospital pathways in which structural cardiac assessment is relevant but not embedded at the point of decision-making, including perioperative pathways, intensive care and non-cardiology specialist services, such as oncology, may face the same hidden dependence on deferred echocardiography. The study was not designed to test those settings directly, but it gives reason to suspect that the problem is broader than is usually acknowledged.

This study has limitations. It was conducted in a single centre and included only 47 patients. The sample size limits precision, particularly in subgroup analyses. The cohort was intentionally enriched for ischaemic disease, which strengthens the clinical relevance of the research question but limits generalisability to broader unselected populations. HFpEF representation was limited, and larger multicentre studies are needed to assess performance in preserved-EF phenotypes and across a wider case mix.

At the same time, the real-world cardiology setting is a strength. The central insight of this study did not arise despite that environment, but because of it. If CHART had performed poorly, the study would simply have remained a failed agreement exercise. Because it performed credibly, the hidden workflow deficit became visible. Therefore, this study opens new roads for exploring the potential use of this AI-powered technology in clinical pathways both in hospital and primary care settings in the future.

## Conclusion

CHART showed clinically credible performance for LVEF estimation and HF stratification, particularly in reduced-EF phenotypes. However, the most important finding of this study was not agreement alone. By performing effectively in a cardiology-based real-world setting, CHART exposed a previously under-recognised workflow deficit: the gap between the nominal availability of echocardiography and timely access to structural cardiac assessment in routine care.

What this study laid bare was that the distance between gold-standard diagnostics and real-world access was not marginal, but clinically meaningful, and visible even within a specialist environment in which that gap might have been assumed to be irrelevant. The significance of CHART therefore lies not only in diagnostic performance metrics, but in making visible a hidden delay in cardiac assessment pathways.

CHART is not a replacement for echocardiography. Its more important implication is that it may offer a practical means of narrowing the otherwise obscured gap between gold-standard capability and timely access to actionable structural and functional cardiac information. Larger studies are now needed to confirm these findings across broader populations and workflows, particularly in HFpEF and in care pathways where the absence of immediate structural assessment continues to shape decisions and outcomes.

## Acknowledgements

The authors thank Marc Bisnaire for valuable contribution to project coordination. The authors also thank Ana Freire and Ana Filipa Ferreira (W4Research) for writing support during preparation of the manuscript.

## Data availability statement

Data are available from the corresponding author upon reasonable request.

## Supplementary Material

**Supplementary Table 1.** Binary performance metrics used for CHART’s validation.

| Abbr. | Metric Name | Description |
| --- | --- | --- |
| F1% | F1 score | $F1 = 2 / (1/PPV + 1/SE)$ |
| K% | Cohens Kappa(17) | $K = (TN+FP)/N * (TN+FN)/N + (FN+TP)/N * (FN+TP)/N$ |
| ACC% | Accuracy | $ACC = (TP+TN) / (P + N)$ |
| AUCp% | Parametric Area Under Curve | ROC curve estimated from SE and SP, and AUC calculated under ROC curve |
| SE% | Sensitivity | $SE = TP / (TP+FN)$ |
| SP% | Specificity | $SP = TN / (TN+FP)$ |
| PPV% | Positive Predictive Value | $PPV = TP / (TP+FP)$ |
| NPV% | Negative Predictive Value | $NPV = TN / (TN+FN)$ |
| PR% | Positive Rate | $PR = (TP + FP) / (TP + FP + TN + FN)$<br>(it is not a performance measure, only for comparison purpose) |
| SEsp90% | Sensitivity at specificity of 90% | Simulated sensitivity when SP=90% estimated by parametric ROC curve |
| RMSE | Root Mean Square Error | $RMSE = \sqrt{\frac{1}{N} \sum_{i=1}^n (x_i - y_i)^2}$ |

*P*: positive test, abnormal; *N*: negative test, normal; *FP*: false positive; *FN*: false negative; *TP*: true positive; *TN*: true negative; $x_i$ : predictor variable (CHART); $\bar{x}$ : average value of *x*; $y_i$ : response variable (ECHO); $\bar{y}$ : average value of *y*; *n*: number of samples

## References

1. Savarese G, Becher PM, Lund LH, et al. Global burden of heart failure: a comprehensive and updated review of epidemiology. Cardiovascular Research 2022; 118: 3272–3287. DOI: 10.1093/cvr/cvac013.

2. Wong CW, Tafuro J, Azam Z, et al. Misdiagnosis of Heart Failure: A Systematic Review of the Literature. Journal of Cardiac Failure 2021; 27: 925–933. DOI: 10.1016/j.cardfail.2021.05.014.

3. Pieske B. Heart failure with preserved ejection fraction—a growing epidemic or ‘The Emperor’s New Clothes?’. European Journal of Heart Failure 2011; 13: 11–13. DOI: 10.1093/eurjhf/hfq215.

4. Sabbah HN. Silent disease progression in clinically stable heart failure. Eur J Heart Fail 2017; 19: 469–478. DOI: 10.1002/ejhf.705.

5. Taylor CJ. Diagnosing heart failure: challenges in primary care. Heart 2019; 105: 663–664. DOI: 10.1136/heartjnl-2018-314396.

6. Magnussen C and Blankenberg S. Biomarkers for heart failure: small molecules with high clinical relevance. J Intern Med 2018; 283: 530–543. DOI: 10.1111/joim.12756.

7. Lund LH, Pitt B and Metra M. Left ventricular ejection fraction as the primary heart failure phenotyping parameter. Eur J Heart Fail 2022; 24: 1158–1161. DOI: 10.1002/ejhf.2576.

8. Otto CM. Heartbeat: Primary care delays in heart failure diagnosis. Heart 2019; 105: 661–662. DOI: 10.1136/heartjnl-2019-315117.

9. Lala A and Desai AS. The role of coronary artery disease in heart failure. Heart Fail Clin 2014; 10: 353–365. DOI: 10.1016/j.hfc.2013.10.002.

10. Cardio-Phoenix. Cardio-HART TM “CHART” Breakthrough cardiac diagnostic system for the early detection of Cardiovascular Diseases, Heart Failure, and Heart Valve diseases. Available at: https://www.cardiophoenix.com/ (accessed 3 July 2024).

11. Nogueira MA, Calcagno S, Campbell N, et al. Detecting heart failure using novel bio-signals and a knowledge enhanced neural network. Computers in Biology and Medicine 2023; 154: 106547. DOI: 10.1016/j.compbiomed.2023.106547.

12. McDonagh TA, Metra M, Adamo M, et al. 2021 ESC Guidelines for the diagnosis and treatment of acute and chronic heart failure. European Heart Journal 2021; 42: 3599–3726. DOI: 10.1093/eurheartj/ehab368.

13. Lang RM, Badano LP, Mor-Avi V, et al. Recommendations for Cardiac Chamber Quantification by Echocardiography in Adults: An Update from the American Society of Echocardiography and the European Association of Cardiovascular Imaging. European Heart Journal - Cardiovascular Imaging 2015; 16: 233–271. DOI: 10.1093/ehjci/jev014.

14. Bland JM and Altman DG. Statistical methods for assessing agreement between two methods of clinical measurement. Lancet 1986; 1: 307–310.

15. Pellikka PA, She L, Holly TA, et al. Variability in Ejection Fraction Measured By Echocardiography, Gated Single-Photon Emission Computed Tomography, and Cardiac Magnetic Resonance in Patients With Coronary Artery Disease and Left Ventricular Dysfunction. JAMA Netw Open 2018; 1: e181456. DOI: 10.1001/jamanetworkopen.2018.1456.

16. Nazir MS, Okafor J, Murphy T, et al. Echocardiography versus Cardiac MRI for Measurement of Left Ventricular Ejection Fraction in Individuals with Cancer and Suspected Cardiotoxicity. Radiology: Cardiothoracic Imaging 2024; 6. DOI: 10.1148/ryct.230048.

17. Bellenger NG, Burgess MI, Ray SG, et al. Comparison of left ventricular ejection fraction and volumes in heart failure by echocardiography, radionuclide ventriculography and cardiovascular magnetic resonance; are they interchangeable? Eur Heart J 2000; 21: 1387–1396. DOI: 10.1053/euhj.2000.2011.

18. Istratoaie S, Gargani L, Popescu BA, et al. How to diagnose heart failure with preserved ejection fraction. European Heart Journal - Cardiovascular Imaging 2024; 25: 1505–1516. DOI: 10.1093/ehjci/jeae183.

19. Danzmann LC, Belyavskiy E, Jorge AJL, et al. The challenge of HFpEF diagnosis in Brazil. ABC Heart Failure & Cardiomyopathy. 2021;1(1). doi:10.36660/abcheartfailure.20210001

20. Beale AL, Meyer P, Marwick TH, et al. Sex Differences in Cardiovascular Pathophysiology. Circulation 2018; 138: 198–205. DOI: 10.1161/CIRCULATIONAHA.118.034271.

21. Lam CSP, Arnott C, Beale AL, et al. Sex differences in heart failure. Eur Heart J 2019; 40: 3859–3868c. DOI: 10.1093/eurheartj/ehz835.

22. Bethge A, Penciu O, Baksh S, et al. Appropriateness vs value: Echocardiography in primary care. Clin Cardiol 2017; 40: 1212–1217. 20171216. DOI: 10.1002/clc.22810.

23. O’Sullivan JW, Albasri A, Nicholson BD, et al. Overtesting and undertesting in primary care: a systematic review and meta-analysis. BMJ Open 2018; 8: e018557. 20180211. DOI: 10.1136/bmjopen-2017-018557.

